# Serotype distribution and risk factors associated with pneumococcal carriage among children with otitis media in Peninsular Malaysia (2023-2025): A cross-sectional study in the early post-vaccination era

**DOI:** 10.64898/2026.07.28.26359181

**Authors:** Chen Zhe Tang, Nurul Hanis Ramzi, Nur Alia Johari, Ailin Razali, Zamzil Amin Asha’Ari, Norhidayah Kamarudin, Azwanis Abdul Hadi, Saraiza Abu Bakar, Khadijah Mohd Nor, Chun Wie Chong, Alex J.J. Lister, David W. Cleary, Stuart C. Clarke, Lokman Hakim Sulaiman

## Abstract

**Background:** Otitis media is a leading cause of childhood morbidity and presents a significant healthcare burden worldwide. *Streptococcus pneumoniae* is a major aetiological agent of OM, and the introduction of pneumococcal conjugate vaccines into the Malaysian National Immunisation Programme in 2020 would have altered pneumococcal carriage and serotype distribution. This study aimed to determine pneumococcal carriage, serotype distribution, and associated risk factors among children with OM in the early post-PCV era in Peninsular Malaysia.

**Methods and Findings:** A total of 360 children with OM were recruited from hospitals on the east and west coasts of Peninsular Malaysia between 2023 and 2025. Nasopharyngeal and middle ear fluid samples were collected for Spn isolation by culture, followed by serotyping using multiplex PCR. Sociodemographic, environmental, and medical history data were analysed for associations with pneumococcal carriage using chi-square, Fisher’s exact tests, and logistic regression. Pneumococcal carriage was detected in 26.7% of children in either NP or MEF samples, with carriage rates of 25.6% and 1.9% in NP and MEF samples, respectively. The most prevalent serotypes were 23A, 15B/15C, non-typable strains, 19F, and 11A/11D. Daycare attendance (p = 0.012, aOR [95% CI]: 2.177 [1.186 – 3.995] and residence in rural areas (p = 0.019, aOR [95% CI]: 2.476 [1.159 – 5.292] were significantly associated with pneumococcal carriage. The main limitation of the study was the reliance on self-reported questionnaire data, which may have introduced recall bias and reporting errors.

**Conclusions:** The predominance of non-vaccine serotypes and non-typable Spn indicates ongoing serotype replacement and the emergence of phase-variant strains in the early post-PCV era. Continued surveillance is essential to monitor these changes and inform the development of next-generation pneumococcal vaccines. This study was registered under clinical trial registration number NCT05429541.

## Introduction

Otitis media (OM), or middle ear infection is among the most common childhood diseases, representing one of the leading causes of paediatric morbidity. Although often self-limiting, OM may lead to serious complications such as hearing loss, impacting speech and language development in critical developmental years [1]. Globally, OM remains a major public health concern, with approximately 390 million cases and an estimated 2.5 million disability-adjusted life years reported annually [2]. In Malaysia, the direct healthcare costs associated with acute OM alone were previously estimated at RM667 million (US$207 million), with additional indirect burdens arising from caregiver work absenteeism and impaired quality of life [3].

Although commonly detected as part of the normal upper respiratory tract microbiota, *Streptococcus pneumoniae*, *Haemophilus influenzae*, and *Moraxella catarrhalis* are important opportunistic pathogens implicated in the development of OM [4]. Young children are particularly susceptible to OM due to their immature immune systems and the anatomical characteristics of their Eustachian tubes, which facilitate the migration of pathogens from the nasopharynx to the middle ear [5]. Among these pathogens, *S. pneumoniae* (Spn) is one of the most commonly identified bacterial causes of OM and has been a major focus of public health interventions due to its numerous serotypes and remarkable genetic adaptability and variability [6,7].

The most effective strategy for preventing pneumococcal disease has been the implementation of pneumococcal vaccination programmes, particularly through the use of pneumococcal conjugate vaccines (PCVs) in early childhood. Although PCVs have long been available through the private healthcare sector in Malaysia, their incorporation into the National Immunisation Programme (NIP) occurred relatively recently, with the inclusion of the ten-valent PCV (PCV10, Synflorix®, GlaxoSmithKline) in 2020 [8]. Subsequently, the thirteen-valent PCV (PCV13, Prevnar 13®, Pfizer) was introduced in 2023 as part of the Catch-up Pneumococcal Vaccination Programme and was later adopted into the routine NIP schedule [9]. These initiatives have achieved excellent vaccine uptake, reaching approximately 99% coverage among eligible children in 2024 [9]. However, post-PCV surveillance data on circulating pneumococcal strains remain limited.

To the best of our knowledge, no studies have comprehensively characterised pneumococcal carriage, serotype distribution, and OM-associated strains in Malaysian children following the introduction of PCVs. Such surveillance is important given the potential for vaccine-driven changes in pneumococcal epidemiology, including shifts in serotype prevalence and the emergence of non-vaccine serotypes (NVTs) [10,11]. In addition, understanding factors associated with pneumococcal carriage is also essential for informing disease prevention strategies. Previous studies have identified a range of environmental, demographic, health-related, and socioeconomic factors that influence pneumococcal colonisation in children [12]. However, investigations specifically focusing on risk factors for pneumococcal carriage among children with OM are limited, especially in the Malaysian setting. Identifying locally relevant risk factors may facilitate the development of targeted public health interventions aimed at reducing pneumococcal transmission and the burden of Spn-associated OM. Thus, this study was conducted to characterise pneumococcal carriage and serotype distribution among children ≤5 years of age with OM in Peninsular Malaysia and to identify factors associated with pneumococcal carriage within this population in the early post-PCV era.

## Methods

### Study population and sample collection

A cross-sectional surveillance study was conducted between April 2023 and August 2025 across two sentinel sites in Peninsular Malaysia: Kuantan, Pahang and Serdang, Selangor, representing the east and west coasts, respectively. A total of 360 paediatric patients were recruited, with 180 participants enrolled from each site. Eligible participants were Malaysian children aged between 3 months and 5 years who were clinically diagnosed with acute otitis media (AOM), otitis media with effusion (OME), or chronic suppurative otitis media (CSOM). Exclusion criteria included the presence of tympanostomy or ventilation tubes, a history of ear, nose and throat (ENT) surgery, or receipt of antibiotics within one month prior to sample collection. Written informed consent was obtained from parents or legally authorised representatives before enrolment.

Clinical specimens were collected at the time of enrolment according to local clinical practice guidelines. One nasopharyngeal (NP) swab and one middle ear fluid (MEF) specimen were obtained from each participant. MEF samples were collected either by swabbing of otorrhoeal discharge or by tympanocentesis when clinically indicated. Following collection, swabs were immediately placed into 1 mL of skim milk-tryptone-glucose-glycerol (STGG) transport medium and stored at −80°C. Samples were transported on dry ice from the sentinel sites to the research laboratory and maintained at −80°C until further processing.

Information on sociodemographic characteristics, environmental exposures, clinical findings, medical history, and vaccination status was collected using a standardised questionnaire administered through face-to-face interviews with the participants’ legally authorised representatives.

### Pneumococcal isolation and serotyping

Spn was identified and isolated from individual NP and MEF specimens by conventional microbiological techniques, and presumptive pneumococcal isolates were identified based on colony morphology and optochin susceptibility testing. Where multiple morphologically distinct pneumococcal colonies were observed within a specimen, each colony type was sub-cultured and retained for further analysis to account for the possibility of co-colonisation by different serotypes. Serotyping was subsequently performed using multiplex polymerase chain reaction (PCR) for 41 serospecificities [13]. Isolates with a detectable *cpsA* band but no corresponding serotype-specific band were classified as non-typable (NT) Spn.

### Statistical analysis

Participant data were analysed using IBM SPSS Statistics version 29 (IBM Corp., USA). Briefly, sociodemographic characteristics, environmental exposure factors, vaccination history, and clinical signs and symptoms were evaluated as potential risk factors for NP Spn carriage. Categorical variables were compared using Pearson’s chi-square test or Fisher’s exact test, as appropriate, while continuous variables were analysed using the Mann-Whitney U test. All statistical tests were two-sided, and a p-value of ≤ 0.05 was considered statistically significant. Variables that were significantly associated with the univariate analysis were entered into the logistic regression model to identify independent predictors of carriage. The regression models were adjusted for potential confounders (recruiting sites, age group and sex) for accurate estimation of the independent effects of the predictors. Adjusted odds ratios (aORs) and corresponding 95% confidence intervals (CIs) were calculated for variables included in the final model.

## Results

### Pneumococcal carriage and serotype distribution

Overall, 26.7% (96/360) of participants were positive for Spn in either NP or MEF samples. Spn was detected in 25.6% (92/360) of NP samples and 1.9% (7/360) of MEF samples, with only three participants yielding positive cultures from both specimen types. The proportion of Spn-positive cases in NP and/or MEF samples was similar across OM subtypes: 26.5% (74/279) AOM, 33.3% (2/6) CSOM, and 26.7% (20/75) OME cases, with the highest detection observed in CSOM cases. A high degree of discordance was observed between NP and MEF cultures, as most Spn-positive NP samples did not correspond to Spn-positive MEF samples. Among the seven Spn-positive MEF samples, only three (42.9%) demonstrated concordant NP carriage of the same serotype, including two serotype 19F isolates and one serotype 11A/11D isolate. No corresponding NP carriage isolate was detected in the remaining four cases, for which the serotypes identified were 23A, 19A, and 35B, as well as one NT isolate.

One participant harboured two distinct serotypes within a single NP specimen, resulting in a total of 93 NP isolates available for serotype analysis (**Fig 1**). Across both sentinel sites, the most frequently identified serotypes were 23A (18.3%, 17/93), followed by 15B/15C (15.1%, 14/93), NT Spn (14.0%, 13/93), 19F (9.7%, 9/93), and 11A/11D (9.7%, 9/93). However, the distribution of Spn serotypes differed between sites. In Serdang (n = 47), the most common serotypes were NT Spn (23.4%, 11/47), 15B/15C (14.9%, 7/47), and 11A/11D (12.8%, 6/47). In contrast, isolates from Kuantan (n = 46) were predominantly characterised by serotypes 23A (28.3%, 13/46), 6A/6B (17.4%, 8/46), 19F (15.2%, 7/46), and 15B/15C (15.2%, 7/46).

**Fig 1:**
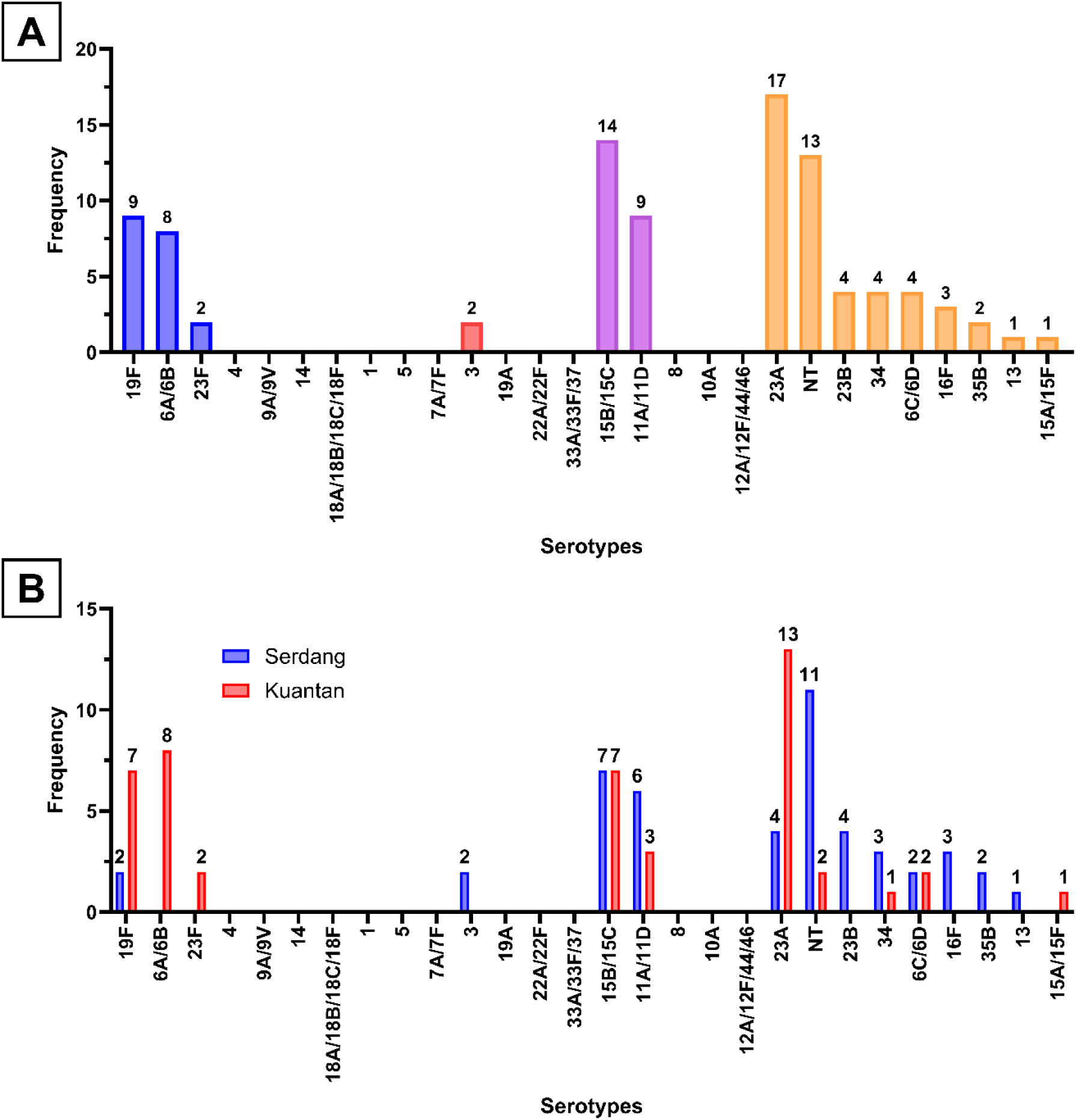
Distribution of pneumococcal serotypes identified via multiplex PCR among children aged ≤5 years with otitis media (n = 93). (A) Overall serotype distribution across both sentinel sites. Colours indicate serotypes covered by PCV10 (blue), PCV13 (red), PCV20 (purple), and non-vaccine serotypes (orange). (B) Site-specific serotype distribution by site: Serdang (n = 47) and Kuantan (n = 46). NT: non-typable Spn. Note: 6A is not included in PCV10; 15C and 11D are not included in PCV20.

The overall estimated vaccine serotype (VT) coverage among NP isolates was 20.4% (19/93) for PCV7 and PCV10, 22.6% (21/93) for PCV13 and PCV15, and 47.3% (44/93) for PCV20. More than half of all isolates (52.7%, 49/93) were classified as NVTs, including NT strains. Among the 93 NP-cultured Spn isolates and four additional MEF-only isolates without corresponding NP detection (n = 97), serotypes covered by PCV10 accounted for 19.6% (19/97), while coverage increased to 22.7% (22/97) for both PCV13 and PCV15, and to 46.4% (45/97) for PCV20. NVTs comprised the majority at 53.6% (52/97). VTs covered by different PCV formulations significantly differed between Serdang and Kuantan (**Table 1** and **Fig 2**). Kuantan consistently exhibited higher proportions of VTs than Serdang across all PCV formulations, whereas NVTs predominated in Serdang and were significantly more prevalent than in Kuantan.

**Fig 2:**
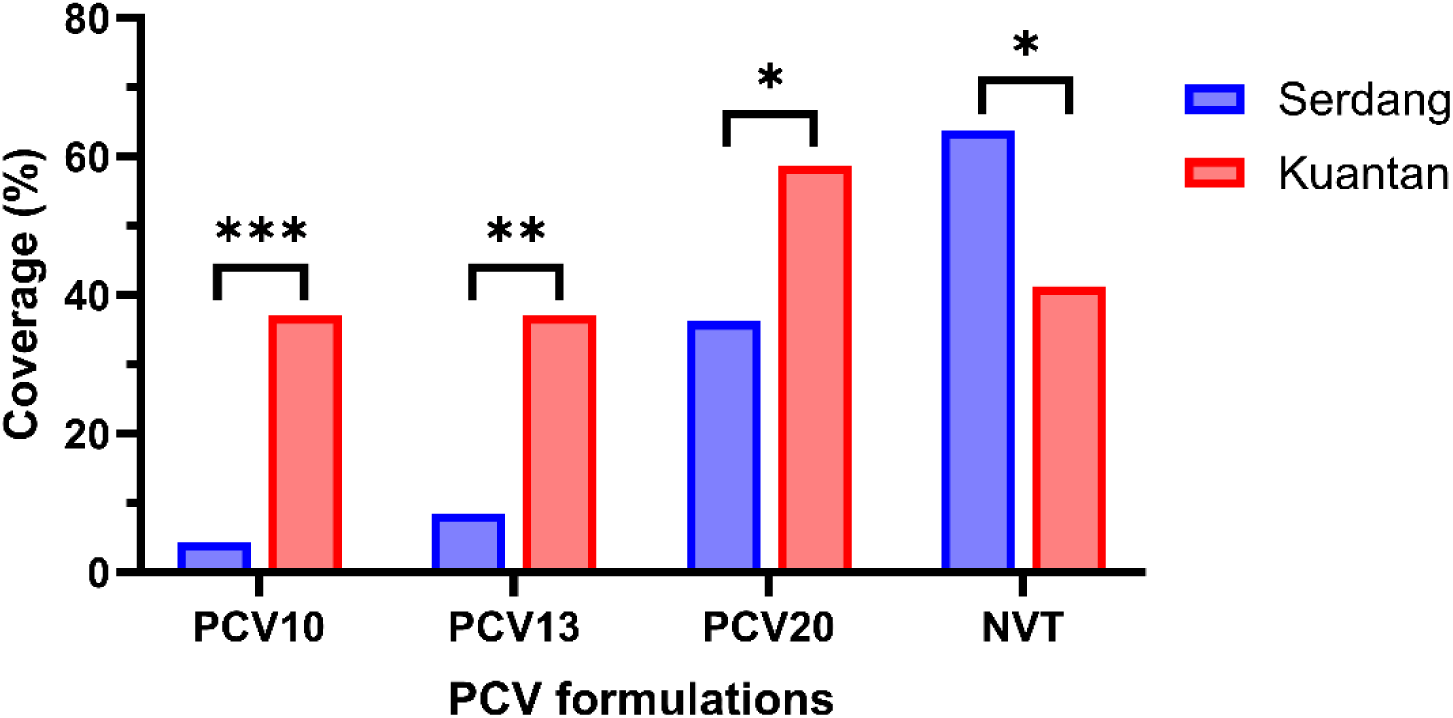
Difference in identified pneumococcal serotypes in nasopharyngeal carriage in OM children across sentinel sites.

**Table 1:** Serotype coverage of pneumococcal conjugate vaccines (PCV7/10, PCV13/15, PCV20) and prevalence of non-vaccine serotypes (NVT) among nasopharyngeal pneumococcal isolates from children with otitis media.

|  | Kuantan (n = 46),<br>n (%) | Serdang (n = 47), n<br>(%) | p-value |
| --- | --- | --- | --- |
| **PCV10** |  |  | **<0.001** |
| Yes | 17 (37.0) | 2 (4.3) |  |
| No | 29 (63.0) | 45 (95.7) |  |
| **PCV13** |  |  | **0.001** |
| Yes | 17 (37.0) | 4 (8.5) |  |
| No | 29 (63.0) | 43 (91.5) |  |
| **PCV20** |  |  | **0.030** |
| Yes | 27 (58.7) | 17 (36.2) |  |
| No | 19 (41.3) | 30 (63.8) |  |
| **NVT** |  |  | **0.030** |
| Yes | 19 (41.3) | 30 (63.8) |  |
| No | 27 (58.7) | 17 (36.2) |  |

### PCV vaccination status and pneumococcal serotypes

21.1% (76/360) of the recruited children were unvaccinated with PCV, 29.7% (107/360) had received one or two primary PCV doses, and 49.2% (177/360) were fully vaccinated. Among the 278 children aged ≥16 months who were eligible to complete the schedule, 114 (41.0%) had not completed the recommended three-dose PCV regimen. No significant association was observed between PCV vaccination status and NP carriage of Spn in the overall analysis (p = 0.213).

VTs persisted in NP carriage included PCV10 serotypes (19F, 6B, and 23F) and PCV13-exclusive serotypes (6A and 3). Serotypes 19F and 6A/6B predominated, accounting for 81.0% (17/21) of PCV13 VT isolates and 18.3% (17/93) of all carriage isolates. Among children carrying 19F and 6A/6B, 55.6% (5/9) and 50.0% (4/8), respectively, were unvaccinated, with most being older than the recommended age for complete vaccination. In contrast, serotype 23A was frequently detected regardless of vaccination status, with 29.4% (5/17) of carriers being unvaccinated and 47.1% (8/17) having completed the three-dose schedule. PCV13 VT serotypes were significantly associated with PCV uptake (p = 0.042) and unvaccinated children constituted a higher proportion of PCV13 VT carriers than non-PCV13 carriers (47.6% [10/21] vs 19.4% [14/72]).

### Sociodemographic risk factors for pneumococcal carriage

**Table 2** summarises the distribution of sociodemographic characteristics, environmental exposure factors, and PCV vaccination status associated with NP pneumococcal carriage in the combined study population from both sentinel sites.

**Table 2:** Sociodemographic, perinatal, environmental exposure, and vaccination characteristics of under-5 children with otitis media by nasopharyngeal pneumococcal carriage status.

| Characteristics | Nasopharyngeal pneumococcal carriage |  | p-value |
| --- | --- | --- | --- |
|  | No (n = 268) | Yes (n = 92) |  |
| Age (months), median (IQR) | 29 (16-47.75) | 30.5 (20.5-47) | 0.557 |
| Age group, n (%) |  |  | 0.706 |
| ≤2yo | 105 (39.2) | 34 (37.0) |  |
| >2yo | 163 (60.8) | 58 (63.0) |  |
| Sex, n (%) |  |  | 0.231 |
| Male | 135 (50.4) | 53 (57.6) |  |
| Female | 133 (49.6) | 39 (42.4) |  |
| Residential area, n (%) |  |  | 0.068 |
| Urban/Suburban | 236 (88.1) | 74 (80.4) |  |
| Rural | 32 (11.9) | 18 (19.6) |  |
| Residence type, n (%)<sup>a</sup> |  |  | 0.423 |
| Landed house* | 200 (74.9) | 65 (70.7) |  |
| Flat/apartment | 67 (25.1) | 27 (29.3) |  |
| Household size, median (IQR)<sup>b</sup> | 5 (4-6) | 4 (3-5) | 0.072 |
| **Number of rooms, median (IQR)<sup>c</sup>** | 3 (3-3) | 3 (3-3) | 0.675 |
| **People-per-room, median (IQR)<sup>c</sup>** | 1.5 (1.26-1.76) | 1.33 (1-1.67) | **0.024** |
| **Number of children in residence, median (IQR)** | 2 (1-2) | 1 (1-2) | 0.260 |
| **Gestation age (weeks), median (IQR)<sup>c</sup>** | 38 (38-39) | 38 (38-39) | 0.437 |
| **Gestational term, n (%)<sup>d</sup>** |  |  | 0.154 |
| Pre-term | 24 (9.1) | 4 (4.4) |  |
| Term | 241 (90.9) | 87 (95.6) |  |
| **Birth weight (g)<sup>e</sup>** | 2970 (2740-3200) | 3000 (2800-3200) | 0.725 |
| **Daycare attendance, n (%)<sup>b</sup>** |  |  | **0.006** |
| Yes | 163 (61.0) | 70 (76.9) |  |
| No | 104 (39.0) | 21 (23.1) |  |
| **Smoke exposure, n (%)<sup>b</sup>** |  |  | 0.238 |
| Yes | 120 (45.1) | 35 (38.0) |  |
| No | 146 (54.9) | 57 (62.0) |  |
| **Breastfeeding status (≤2yo only), n (%)<sup>f</sup>** |  |  | 0.645 |
| Breast exclusively | 30 (28.8) | 12 (35.3) |  |
| Mixed feeding | 45 (43.3) | 15 (44.1) |  |
| No breast milk | 29 (27.9) | 7 (20.6) |  |
| **PCV vaccination status, n (%)** |  |  | 0.213 |
| None taken | 53 (19.8) | 23 (25.0) |  |
| 1 to 2 primary doses | 86 (32.1) | 21 (22.8) |  |
| 2 primary + booster | 129 (48.1) | 48 (52.2) |  |

\*Landed house: non-high-rise building including terrace, semi-detached house, bungalow, etc.
<sup>a</sup>Residence type missing data n=1/360
<sup>b</sup>Household size, daycare attendance, and smoke exposure missing data n=2/360
<sup>c</sup>Number of rooms, people-per-room, and gestational age missing data n=6/360
<sup>d</sup>Gestational term missing data n=4/360
<sup>e</sup>Birth weight missing data n=3/360
<sup>f</sup>Breastfeeding status (≤2yo only) missing data n=1/139.

Among the sociodemographic and environmental exposure factors analysed across both sentinel sites, only daycare attendance and people-per-room were significantly associated with NP carriage of Spn (p = 0.006 and p = 0.024) respectively.

The two sentinel sites differed significantly in several sociodemographic and environmental exposure characteristics, including age, sex, residential area, residence type, daycare attendance, household size, and people-per-room (Table 3). Despite these differences, the prevalence of NP carriage of Spn was comparable between Kuantan (25.0%, n = 45/180) and Serdang (26.1%, n = 47/180), with no significant difference observed between sites (p = 0.809).

**Table 3:** Sociodemographic, perinatal, environmental exposure, and vaccination characteristics of under-5 children with otitis media by sentinel sites.

| Characteristics | Kuantan (n = 180) | Serdang (n = 180) | p-value |
| --- | --- | --- | --- |
| **Age (months), median (IQR)** | 35.50 (20.50 – 48.75) | 28.00 (15.00 – 45.00) | **0.022** |
| **Age group, n (%)** |  |  | **0.040** |
| ≤2yo | 60 (33.3) | 79 (43.9) |  |
| >2yo | 120 (66.7) | 101 (56.1) |  |
| **Sex, n (%)** |  |  | **0.006** |
| Male | 81 (45.0) | 107 (59.4) |  |
| Female | 99 (55.0) | 73 (40.6) |  |
| **Residential area, n (%)** |  |  | **<0.001** |
| Urban/Suburban | 135 (75.0) | 175 (97.2) |  |
| Rural | 45 (25.0) | 5 (2.8) |  |
| **Residence type, n (%)<sup>a</sup>** |  |  | **<0.001** |
| Landed house* | 175 (97.8) | 90 (50.0) |  |
| Flat/apartment | 4 (2.2) | 90 (50.0) |  |
| **Household size, median (IQR)<sup>b</sup>** | 5.00 (4.00 – 6.00) | 4.00 (4.00 – 5.00) | **<0.001** |
| **Number of rooms, median (IQR)<sup>c</sup>** | 3.00 (3.00 – 4.00) | 3.00 (3.00 – 3.00) | 0.104 |
| **People-per-room, median (IQR)<sup>c</sup>** | 1.66 (1.33 – 2.00) | 1.33 (1.00 – 1.67) | **<0.001** |
| **Number of children in residence, median (IQR)** | 1.00 (1.00 – 2.00) | 2.00 (1.00 – 2.00) | 0.462 |
| **Gestation age (weeks), median (IQR)<sup>c</sup>** | 38.00 (37.75 – 40.00) | 38.00 (38.00 – 39.00) | 0.585 |
| **Gestational term, n (%)<sup>d</sup>** |  |  | 0.396 |
| Pre-term | 16 (9.1) | 12 (6.7) |  |
| Term | 160 (90.9) | 168 (93.3) |  |
| **Birth weight (g), median (IQR)<sup>e</sup>** | 3000 (2660 – 3365) | 2950 (2800 – 3200) | 0.439 |
| **Daycare attendance, n (%)<sup>b</sup>** |  |  | **<0.001** |
| Yes | 93 (52.2) | 140 (77.8) |  |
| No | 85 (47.8) | 40 (22.2) |  |
| **Smoke exposure, n (%)<sup>b</sup>** |  |  | 0.293 |
| Yes | 82 (46.1) | 73 (40.6) |  |
| No | 96 (53.9) | 107 (59.4) |  |
| **Breastfeeding status ($\leq 2$yo only), n (%)<sup>f</sup>** |  |  | 0.086 |
| Breast exclusively | 15 (25.4) | 27 (34.2) |  |
| Mixed feeding | 23 (39.0) | 37 (46.8) |  |
| No breast milk | 21 (35.6) | 15 (19.0) |  |
| **PCV vaccination status, n (%)** |  |  | **<0.001** |
| None taken | 55 (30.6) | 21 (11.7) |  |
| 1 to 2 primary doses | 57 (31.7) | 50 (27.8) |  |
| 2 primary + booster | 68 (37.8) | 109 (60.6) |  |

\*Landed house: non-high-rise building including terrace, semi-detached house, bungalow, etc.
<sup>a</sup>Residence type missing data n=1/360
<sup>b</sup>Household size, daycare attendance, and smoke exposure missing data n=2/360
<sup>c</sup>Number of rooms, people-per-room, and gestational age missing data n=6/360
<sup>d</sup>Gestational term missing data n=4/360
<sup>e</sup>Birth weight missing data n=3/360
<sup>f</sup>Breastfeeding status ( $\leq 2$ yo only) missing data n=1/139.

PCV uptake differed significantly between the two sentinel sites, with a higher proportion of children from Serdang (60.6%) completing the primary series and booster dose compared with those from Kuantan (37.8%) (**Table 3**). Logistic regression analyses were adjusted for site, age group and sex to account for potential differences in participant characteristics associated with pneumococcal carriage (**Table 4**).

**Table 4:** Risk factors associated with nasopharyngeal pneumococcal carriage among under-5 children with otitis media.

| Characteristics | Category | aOR (95% CI) | p-value |
| --- | --- | --- | --- |
| Residential area | Urban/suburban (reference) | 2.476 (1.159 – 5.292) | **0.019** |
|  | Rural |  |  |
| Residence type | Landed house (reference) | 1.393 (0.717 – 2.706) | 0.328 |
|  | Flat/apartment |  |  |
| People-per-room | N/A (continuous) | 0.501 (0.286 – 0.879) | 0.016 |
| Daycare attendance | No (reference) | 2.177 (1.186 – 3.995) | **0.012** |
|  | Yes |  |  |
| PCV vaccination status | 2 primary + booster (reference) | - | Overall: 0.102 |
|  | None taken | 0 dose vs 3 dose: 1.308 (0.678 – 2.525) | 0.423 |
|  | 1 to 2 primary doses | 1-2 dose vs 3 dose: 0.548 (0.278 – 1.078) | 0.081 |

Following adjustment, residential area was significantly associated with pneumococcal carriage, as were the daycare attendance and people-per-room. Children residing in rural areas had approximately 2.5 times higher odds of pneumococcal carriage than those living in urban/suburban areas (aOR = 2.476, 95% CI: 1.159 – 5.292, p = 0.019). Similarly, children who had attended daycare during the month prior to sampling had approximately twice the odds of pneumococcal carriage compared with those who had not (aOR = 2.177, 95% CI: 1.186 – 3.995, p = 0.012). In contrast, increasing people-per-room was associated with lower odds of pneumococcal carriage, with each one-unit increase in people-per-room corresponding to a 49.9% reduction (aOR = 0.501, 95% CI: 0.286 – 0.879, p = 0.016).

### Association of OM clinical characteristics with pneumococcal carriage

None of the clinical signs or clinical history variables were significantly associated with NP carriage of Spn. Of the symptoms evaluated, only hearing difficulty was significantly associated with carriage (p = 0.019), with affected children having 2.53 times higher odds of carriage than those without (OR = 2.53, 95% CI: 1.137–5.629). Nasal/ear discharge was marginally associated with carriage (p = 0.057), with an inverse relationship observed (OR = 0.615, 95% CI: 0.372–1.017).

Furthermore, no significant association was observed between OM subtype and NP pneumococcal carriage (p = 0.931). Although OM subtype distribution differed significantly between sentinel sites (p = 0.032), with Serdang showing a higher proportion of AOM cases than Kuantan (OR = 1.730, 95% CI: 1.045–2.863), adjustment for study site did not alter the findings. OM subtype remained not significantly associated with pneumococcal carriage (p = 0.908), with no significant differences observed between children with AOM and those with OME/CSOM.

## Discussion

The present study found that 25.6% of children with OM carried Spn in the nasopharynx, comparable to those reported in recent paediatric surveillance studies conducted in Peninsular Malaysia (15.4–39.6%) [9,14,15]. However, the limited local pre-PCV carriage data precludes the reliable assessment of changes in carriage prevalence following nationwide PCV implementation. Compared with neighbouring countries of similar socioeconomic status, such as Indonesia, the carriage prevalence observed in Malaysia appears to be considerably lower [16,17].

Moreover, the pneumococcal serotypes identified were broadly consistent with those reported in previous OM studies. Serotypes such as 19F and 19A remain common causes of disease, whereas detection of non-vaccine serotypes, including 23A and 35B, may reflect the increasing contribution of non-PCV serotypes in the post-vaccination era [18,19]. Although the small number of Spn-positive MEF samples here limits definitive conclusions, these findings support the evolving epidemiology of pneumococcal OM and the need for ongoing serotype surveillance.

Furthermore, our study also observed discordance between NP and MEF pneumococcal detection, with only 42.9% concordance in serotype identification. This is consistent with published data showing 40.5–58.3% identical serotype recovery from both sites [20,21]. The discordance reflects higher pneumococcal serotype diversity in NP colonisation compared to the middle ear [22], indicating that only select serotypes possess sufficient virulence to cause invasive infection. In chronic OM, persistent NP carriage is a recognised risk factor for recurrence [21], and serotypes capable of causing disease may change over time with antimicrobial selection pressure [23]. These findings suggest that serotype-specific factors, rather than simple NP carriage-to-disease concordance, are key determinants of progression to chronic OM.

As tympanocentesis is not routinely performed in AOM management, most MEF specimens were obtained by swabbing the ear canal near the tympanic membrane, potentially limiting their accuracy in representing middle ear microbiology. Nevertheless, NP specimens remain a useful surrogate for investigating OM microbiology and pathogenesis at the population level [24–26], although they cannot establish causality in individual episodes.

A systematic review of pre-PCV Malaysian studies identified serotypes 19F, 23F, 6A, 19A, and 6B as the most common carriage serotypes, with 19F also the predominating in non-invasive pneumococcal disease (non-IPD), accounting for 21.5% of isolates [27]. In contrast, NT pneumococci constituted only a small proportion of isolates in the pre-PCV era (6.3% overall). Yet, despite PCV introduction, VTs 19F and 6A/6B persisted in this study, likely reflecting the sustained colonisation advantage of historically dominant serotypes rather than recent emergence. The VTs included in PCV formulations were originally selected due of their high invasive potential and disease burden in the pre-PCV era [28]. Thus, the continued circulation of these serotypes in the post-PCV era is concerning, particularly 19F and 6A/6B, which are frequently associated with antimicrobial resistance (AMR) [29,30], potentially limiting contributing to the persistence and spread of resistant pneumococcal strains.

Persistence of VTs despite high vaccine uptake has been reported worldwide [31,32], and may reflect vaccine- or antibiotic-resistant variants, reduced niche competition, and serotype-specific immune responses [33]. Serotype 19F is of particular concern, as it frequently persists in the post-PCV era and has been associated with apparent vaccine failure despite its inclusion in current PCVs. Despite 78.9% (284/360) of children having received at least one PCV dose, the continued prominence of 19F suggests that herd protection against this serotype may be incomplete.

One possible explanation is that serotype 19F is inherently less susceptible to antibody-mediated immunity, requiring higher antibody concentrations for effective opsonophagocytic killing than other serotypes [33–35]. Although an IgG concentration of 0.35 µg/mL was the established correlate of protection against IPD for PCV licensure [36,37], subsequent studies have demonstrated that protective thresholds vary substantially between serotypes, and this cutoff does not provide equivalent protection against all pneumococci [18,37,38].

It is also important to recognise that systemic immunity against IPD does not necessarily translate to mucosal protection against OM. While PCVs effectively prevent IPD by inducing circulating IgG antibodies, their ability to prevent NP colonisation is limited as antibody concentrations at mucosal surfaces are substantially lower than in serum [18]. Voysey et al. [39] showed that protection against pneumococcal carriage requires higher antibody concentrations than protection against IPD, with estimated correlates of protection of 0.50 µg/mL for serotype 6B and 2.54 µg/mL for serotype 19F, both exceeding the commonly cited 0.35 µg/mL threshold. Consequently, PCVs are unlikely to eliminate transient colonisation but instead reduce colonisation density [40], thereby lowering disease progression risk and transmission and contributing to herd protection [41].

Furthermore, a recent study demonstrated that most children who received a single dose of PCV10 achieved serotype 6B-specific IgG concentrations above the 0.35 µg/mL threshold but exhibited low opsonophagocytic activity, with similar findings for cross-reactive serotype 6A [42]. However, subsequent primary and booster doses have been shown to markedly improve both vaccine-induced antibody concentration and functionality [35,43,44]. With only 59.0% of children old enough to complete the Malaysian PCV schedule (≥16 months) had received the recommended 2p+1 regimen, the level of protection may have been short of expectations, especially against serotypes requiring higher antibody titres for effective immunity. This incomplete vaccination coverage could therefore have facilitated the persistence of VTs in our cohort.

This finding thus highlights the importance of completing the recommended PCV schedule, particularly the booster dose, as primary doses alone may not induce sufficient or durable immunity. Indeed, some countries that implemented a 3p+0 schedule including Malawi and Australia without a booster have reported suboptimal control of VT-associated diseases [37,45]. More recently, several countries have explored reduced-dose schedules (1p+1) to improve cost-effectiveness [46–49]. These schedules have shown comparable effectiveness against IPD, with no increase in paediatric IPD observed after the UK transitioned from a 2p+1 schedule in 2020 [50,51]. Although a single priming dose induces lower initial immunity than two, booster responses were similar for most serotypes across both schedules [52], suggesting that booster immunogenicity is largely independent of the number of priming doses received. Nevertheless, further studies are needed to determine whether reduced-dose schedules provide non-inferior protection against carriage and mucosal diseases.

Unfortunately, we lacked information on the specific PCV formulations received by individual participants, and multiplex PCR could not distinguish between serotypes 6A and 6B. Given that 6B is included in both PCV10 and PCV13 whereas 6A is only covered by PCV13, this limitation may have masked differences in VT carriage attributable to the type of PCV administered.

Despite the persistence of some VTs, the predominance of NVTs and NT Spn in this study reflects successful PCV implementation and ongoing serotype replacement, consistent with the global trend [53–55]. The dominant NVTs were similar to those recently reported among healthy children in the Klang Valley, Malaysia [15], supporting the occurrence of non-IPDs such as OM is proportional to NP carriage prevalence [56]. The high proportion of NT pneumococci has similarly been observed in other post-PCV settings [55,57].

Although serotypes 23A, 15B/15C, and 11A/11D have traditionally been mainly associated with carriage and non-IPDs [58], they are increasingly reported in IPD and linked to AMR and multidrug resistance (MDR) in the post-PCV era [59–61]. Likewise, NT Spn are emerging as important reservoirs of resistance, with MDR reported in over 25% of isolates globally [62], and AMR observed in 83.3% of NT isolates in Malaysia [15].

Plus, NT pneumococci (i.e., non-encapsulated Spn) are not targeted by current PCVs and their increasing prevalence and involvement in pneumococcal disease following widespread PCV implementation thus pose an emerging public health concern. To overcome limitations associated with serotype replacement and carrier suppression associated with increasing PCV valency, recent efforts have focused on developing universal “pan-pneumococcal” vaccines targeting conserved protein antigens [63].

Malaysia approved twenty-valent PCV (PCV20, Prevnar 20®, Pfizer) in 2025 and recently announced its replacement of PCV13 in the NIP from June 2026 [64,65]. In this cohort, PCV20 would have increased theoretical serotype coverage by 24.7%, mainly through the inclusion of serotypes 15B and 11A. Given its non-inferior immunogenicity and safety profile comparable to earlier PCVs [66–68], PCV20 represents a promising advancement to pneumococcal disease prevention in Malaysia.

Risk factors for pneumococcal carriage were also investigated. Marked differences in sociodemographic characteristics observed between sentinel sites likely reflect variations in local culture and population characteristics. Compared with children from Kuantan, those from Serdang were more likely to reside in urban areas and high-rise housing, have smaller and less crowded households, attend daycare, and adhere to PCV vaccination schedules. After adjusting for sentinel site as a potential confounder, only residing in rural areas and daycare attendance were independently associated with pneumococcal carriage, consistent with previous studies [12].

Residential area became significantly associated with NP carriage only after adjusting for sentinel site, suggesting confounding in the crude analysis. Nevertheless, this finding aligns with previous studies linking rural residence to factors that facilitate pneumococcal transmission, including lower socioeconomic status, crowding, lower vaccine uptake, and smoke exposure [12,69–71]. The significant difference in PCV uptake between sites may partly explain this association. Although smoke exposure was not associated with either residential area (p = 0.181) nor carriage in this study, this may reflect under-reporting given its well-established role in impairing mucosal defences and promoting colonisation [72].

Unexpectedly, household crowding was inversely associated with pneumococcal carriage, contrary to previous reports [73,74]. One possible explanation is that crowding may favour colonisation by competing upper respiratory tract microbes, reducing pneumococcal detection. While an inverse relationship with *Staphylococcus aureus* has been reported [75,76], Spn often co-colonise with species including *H. influenzae* and *M. catarrhalis* [77,78]. Further studies on nasopharyngeal co-colonisation are therefore needed to clarify these findings.

Moreover, daycare attendance is a well-established independent risk factor for pneumococcal carriage. Consistent with previous studies reporting a 1.5- to three-fold greater risk (OR: 1.5– 2.99) [79–82], this association likely reflects increased transmission through close contact, exposure to respiratory secretions, and co-circulating respiratory pathogens that facilitate colonisation [12,83,84].

Several limitations should be considered. The absence of a healthy control group prevented comparisons with children without OM, and only Spn was investigated, without evaluating other otopathogens or co-colonisation. Also, questionnaire-based variables were subject to recall bias and potential misreporting. Future studies should incorporate whole-genome sequencing to characterise pneumococcal lineages and AMR determinants, investigate polymicrobial interactions in OM, and continue surveillance following Malaysia’s transition to PCV20 to monitor vaccine impact.

## Conclusion

Serotype replacement has become evident within five years of the introduction of PCVs into the Malaysian NIP, although VTs such as 19F and 6A/6B continue to persist in carriage in children with OM. The emergence of NVTs, particularly 23A as the predominant serotype followed by 15B/15C and NT Spn, highlights the dynamic changes in the pneumococcal population in the post-PCV era. Daycare attendance and residence in rural areas were identified as significant risk factors associated with pneumococcal carriage. Future studies should further characterise the AMR profiles and genetic lineages of these emerging serotypes to better understand their clinical and public health implications. Given Malaysia’s recent transition to PCV20 in the NIP, continued surveillance of pneumococcal carriage, serotype distribution, and resistance patterns remains crucial to monitor vaccine impact and guide future vaccination strategies.

## Data Availability

The datasets generated and analysed during the current study are not publicly available due to privacy or ethical restrictions.

## Ethics statement

This study was approved by the IMU University Joint Committee on Research and Ethics (IMU-JC) (IRB Ref no.: 4.19/JCM-247/2022) and the Malaysian National Medical Research Register (NMR) (NMRR ID: 22-02418-ACL). Written informed consent was obtained from the parents or legal guardians of all participating children prior to enrolment.

## Abbreviations

AOM: Acute otitis media
CSOM: Chronic suppurative otitis media
IPD: Invasive pneumococcal disease
MEF: Middle ear fluid
NIP: National immunisation programme
NP: Nasopharyngeal
NT: Non-typable
NVT: Non-vaccine type
OM: Otitis media
OME: Otitis media with effusion
PCV: Pneumococcal conjugate vaccine
Spn: *Streptococcus pneumoniae*
VT: Vaccine type

## References

1. Mahadevan M, Navarro-Locsin G, Tan HKK, Yamanaka N, Sonsuwan N, Wang P-C, et al. A review of the burden of disease due to otitis media in the Asia-Pacific. Int J Pediatr Otorhinolaryngol. 2012;76: 623–635. doi:10.1016/j.ijporl.2012.02.031

2. Huang G-J, Lin B-R, Li P-S, Tang N, Fan Z-J, Lu B-Q. The global burden of otitis media in 204 countries and territories from 1992 to 2021: a systematic analysis for the Global Burden of Disease study 2021. Front Public Health. 2025;12: 1519623. doi:10.3389/fpubh.2024.1519623

3. Aljunid S, Maimaiti N, Ahmed Z, Muhammad Nur A, Md Isa Z, Azmi S, et al. Economic Impact of Pneumococcal Protein-D Conjugate Vaccine (PHiD-CV) on the Malaysian National Immunization Programme. Value in Health Regional Issues. 2014;3: 146–155. doi:10.1016/j.vhri.2014.04.008

4. Kim HO, Ha S, Lee SH, Oh YJ, Lee JM, Kim Y-J, et al. Prevalence and Global Distribution of Bacterial Species Associated with Acute Otitis Media in Children: Systematic Review and Meta-Analysis. Antibiotics. 2026;15: 463. doi:10.3390/antibiotics15050463

5. Goulioumis AK, Gkorpa M, Athanasopoulos M, Athanasopoulos I, Gyftopoulos K. The Eustachian Tube Dysfunction in Children: Anatomical Considerations and Current Trends in Invasive Therapeutic Approaches. Cureus. 2022;14: e27193. doi:10.7759/cureus.27193

6. Li J, Zhang J-R. Phase Variation of Streptococcus pneumoniae. Microbiology Spectrum. 2019;7: 10.1128/microbiolspec.gpp3-0005–2018. doi:10.1128/microbiolspec.gpp3-0005-2018

7. Sunmonu GT, Lo SW, Sheppard AE, Ogunniyi AD. Serotype replacement and mobile genetic elements in Streptococcus pneumoniae: a systematic review. Microb Genom. 2025;11: 001497. doi:10.1099/mgen.0.001497

8. Lister AJJ, Dombay E, Cleary DW, Sulaiman LH, Clarke SC. A brief history of and future prospects for pneumococcal vaccination in Malaysia. Pneumonia. 2023;15: 12. doi:10.1186/s41479-023-00114-8

9. Gunasegaran H, Ramzi NH, Hoong Tan AC, Johari NA, Nathan AM, Sulaiman NA, et al. Prevalence of pneumococcal carriage and risk factors for pneumonia and carriage among under-5 children in Malaysia: findings from the MY-Pneumo study. Pneumonia (Nathan). 2025;17: 24. doi:10.1186/s41479-025-00177-9

10. Dhawale P, Shah S, Sharma K, Sikriwal D, Kumar V, Bhagawati A, et al. Streptococcus pneumoniae serotype distribution in low- and middle-income countries of South Asia: Do we need to revisit the pneumococcal vaccine strategy? Human Vaccines & Immunotherapeutics. 2025;21: 2461844. doi:10.1080/21645515.2025.2461844

11. Hanquet G, Krizova P, Dalby T, Ladhani SN, Nuorti JP, Danis K, et al. Serotype Replacement after Introduction of 10-Valent and 13-Valent Pneumococcal Conjugate Vaccines in 10 Countries, Europe. Emerg Infect Dis. 2022;28: 127–138. doi:10.3201/eid2801.210734

12. Neal EFG, Chan J, Nguyen CD, Russell FM. Factors associated with pneumococcal nasopharyngeal carriage: A systematic review. PLOS Global Public Health. 2022;2: e0000327. doi:10.1371/journal.pgph.0000327

13. Centers of Disease Control and Prevention. Streptococcus pneumoniae Detection and Serotyping Using PCR. In: Streptococcus pneumoniae Detection and Serotyping Using PCR [Internet]. 2024 [cited 18 June 2026]. Available: https://www.cdc.gov/strep-lab/php/pneumococcus/serotyping-using-pcr.html

14. Arushothy R, Tan CN, Nor Amdan NA, Mohd Ali MR, Mohd Tap R, Paliappan PA, et al. Serotype diversity and risk factors for pneumococcal carriage among healthy children in Klang Valley, Malaysia: A pre-vaccination cross-sectional study. IJID Regions. 2026;18: 100814. doi:10.1016/j.ijregi.2025.100814

15. Mohamad N, Yahaya MAM, Muthanna A, Desa MNM, Ismail EN, Dapari R, et al. Nasopharyngeal carriage, serotype, antimicrobial susceptibility, and genotype of pneumococci in young healthy children attending daycare centres in Klang Valley, Malaysia. Journal of Global Antimicrobial Resistance. 2025;44: 64–68. doi:10.1016/j.jgar.2025.05.017

16. Daningrat WOD, Amalia H, Ayu IM, Satzke C, Safari D. Carriage of *Streptococcus pneumoniae* in children under five years of age prior to pneumococcal vaccine introduction in Southeast Asia: A systematic review and meta-analysis (2001–2019). Journal of Microbiology, Immunology and Infection. 2022;55: 6–17. doi:10.1016/j.jmii.2021.08.002

17. Kartasasmita CB, Rezeki Hadinegoro S, Kurniati N, Triasih R, Halim C, Gamil A. Epidemiology, Nasopharyngeal Carriage, Serotype Prevalence, and Antibiotic Resistance of Streptococcus pneumoniae in Indonesia. Infect Dis Ther. 2020;9: 723–736. doi:10.1007/s40121-020-00330-5

18. Pichichero M, Malley R, Kaur R, Zagursky R, Anderson P. Acute otitis media pneumococcal disease burden and nasopharyngeal colonization in children due to serotypes included and not included in current and new pneumococcal conjugate vaccines. Expert Review of Vaccines. 2023;22: 118–138. doi:10.1080/14760584.2023.2162506

19. Bhuiyan MU, Hernandez-Suarez G, Pilishvili T, Mayer B, Abbing-Karahagopian V. Incidence and bacterial etiology of acute otitis media in children in the pneumococcal conjugate vaccine era: A systematic literature review and meta-analysis. Vaccine. 2026;75: 128270. doi:10.1016/j.vaccine.2026.128270

20. Cilveti R, Olmo M, Pérez-Jove J, Picazo J-J, Arimany J-L, Mora E, et al. Epidemiology of Otitis Media with Spontaneous Perforation of the Tympanic Membrane in Young Children and Association with Bacterial Nasopharyngeal Carriage, Recurrences and Pneumococcal Vaccination in Catalonia, Spain - The Prospective HERMES Study. PLOS ONE. 2017;12: e0170316. doi:10.1371/journal.pone.0170316

21. Korona-Glowniak I, Zychowski P, Siwiec R, Mazur E, Niedzielska G, Malm A. Resistant Streptococcus pneumoniae strains in children with acute otitis media-high risk of persistent colonization after treatment. BMC Infect Dis. 2018;18: 478. doi:10.1186/s12879-018-3398-9

22. Lewnard JA, Givon-Lavi N, Tähtinen PA, Dagan R. Pneumococcal Phenotype and Interaction with Nontypeable Haemophilus influenzae as Determinants of Otitis Media Progression. Infect Immun. 2018;86: e00727–17. doi:10.1128/IAI.00727-17

23. Fuji N, Pichichero M, Ehrlich RL, Mell JC, Ehrlich GD, Kaur R. Transition of Serotype 35B Pneumococci From Commensal to Prevalent Virulent Strain in Children. Front Cell Infect Microbiol. 2021;11: 744742. doi:10.3389/fcimb.2021.744742

24. Alexandrova AS, Boyanov VS, Gergova RT. Bacterial Profile, Molecular Serotyping, and Key Genetic Determinants for Adhesion, Immune Evasion, and Tissue Spread Among Bulgarian Children with Acute Otitis Media. Genes. 2025;16: 1512. doi:10.3390/genes16121512

25. Ramasubramanian S, Dhanaseelan S, Prateep A, Sivaramakrishnan SA. Common Bacteria Causing Otitis Media in Patients Attending a Tertiary Care Hospital: A Cross-sectional Study. Apollo Medicine. 2025; 09760016251336532. doi:10.1177/09760016251336532

26. Taylor SL, Papanicolas LE, Richards A, Ababor F, Kang WX, Choo JM, et al. Ear microbiota and middle ear disease: a longitudinal pilot study of Aboriginal children in a remote south Australian setting. BMC Microbiol. 2022;22: 24. doi:10.1186/s12866-022-02436-x

27. Lister AJJ, Le CF, Cheah ESG, Desa MNM, Cleary DW, Clarke SC. Serotype distribution of invasive, non-invasive and carried Streptococcus pneumoniae in Malaysia: a meta-analysis. Pneumonia. 2021;13: 9. doi:10.1186/s41479-021-00086-7

28. Cherian T. WHO expert consultation on serotype composition of pneumococcal conjugate vaccines for use in resource-poor developing countries, 26–27 October 2006, Geneva. Vaccine. 2007;25: 6557–6564. doi:10.1016/j.vaccine.2007.06.044

29. Andrejko K, Ratnasiri B, Lewnard JA. Association of Pneumococcal Serotype With Susceptibility to Antimicrobial Drugs: A Systematic Review and Meta-analysis. Clin Infect Dis. 2022;75: 131–140. doi:10.1093/cid/ciab852

30. Sandoval MM, Ruvinsky S, Palermo MC, Alconada T, Brizuela ME, Wierzbicki ER, et al. Antimicrobial resistance of Streptococcus pneumoniae from invasive pneumococcal diseases in Latin American countries: a systematic review and meta-analysis. Front Public Health. 2024;12. doi:10.3389/fpubh.2024.1337276

31. Domai FM, Shrestha D, Shrestha RK, Thimi M, Ntiamoah DO, Hayashi Y, et al. Non-Vaccine Serotype Replacement and Subdominant Persistence of Vaccine Types in Nepalese Infants Following PCV10 Introduction. Vaccines. 2026;14: 73. doi:10.3390/vaccines14010073

32. Grant LR, Sutcliffe CG, Littlepage S, Alexander-Parrish R, Becenti L, Isturiz RE, et al. Persistence of Vaccine Serotype Carriage and Differences in Pneumococcal Carriage by Laboratory Method and Sample Type in Indigenous Individuals in the Southwest United States. J Infect Dis. 2025;231: e1045–e1056. doi:10.1093/infdis/jiaf091

33. Thindwa D, Carcamo PM, Dagan R, Weinberger DM. What is driving the resurgence and persistence of vaccine-targeted pneumococcal serotypes?—Serotype 19F as the paradigm. PLOS Pathogens. 2026;22: e1014065. doi:10.1371/journal.ppat.1014065

34. Melin M, Jarva H, Siira L, Meri S, Käyhty H, Väkeväinen M. Streptococcus pneumoniae Capsular Serotype 19F Is More Resistant to C3 Deposition and Less Sensitive to Opsonophagocytosis than Serotype 6B. Infect Immun. 2009;77: 676–684. doi:10.1128/IAI.01186-08

35. Song JY, Moseley MA, Burton RL, Nahm MH. Pneumococcal Vaccine and Opsonic Pneumococcal Antibody. J Infect Chemother. 2013;19: 412–425. doi:10.1007/s10156-013-0601-1

36. Siber GR, Chang I, Baker S, Fernsten P, O’Brien KL, Santosham M, et al. Estimating the protective concentration of anti-pneumococcal capsular polysaccharide antibodies. Vaccine. 2007;25: 3816–3826. doi:10.1016/j.vaccine.2007.01.119

37. Swarthout TD, Henrion MYR, Thindwa D, Meiring JE, Mbewe M, Kalizang’Oma A, et al. Waning of antibody levels induced by a 13-valent pneumococcal conjugate vaccine, using a 3 + 0 schedule, within the first year of life among children younger than 5 years in Blantyre, Malawi: an observational, population-level, serosurveillance study. The Lancet Infectious Diseases. 2022;22: 1737–1747. doi:10.1016/S1473-3099(22)00438-8

38. Andrews NJ, Waight PA, Burbidge P, Pearce E, Roalfe L, Zancolli M, et al. Serotype-specific effectiveness and correlates of protection for the 13-valent pneumococcal conjugate vaccine: a postlicensure indirect cohort study. The Lancet Infectious Diseases. 2014;14: 839–846. doi:10.1016/S1473-3099(14)70822-9

39. Voysey M, Fanshawe TR, Kelly DF, O’Brien KL, Kandasamy R, Shrestha S, et al. Serotype-Specific Correlates of Protection for Pneumococcal Carriage: An Analysis of Immunity in 19 Countries. Clin Infect Dis. 2018;66: 913–920. doi:10.1093/cid/cix895

40. German EL, Solórzano C, Sunny S, Dunne F, Gritzfeld JF, Mitsi E, et al. Protective effect of PCV vaccine against experimental pneumococcal challenge in adults is primarily mediated by controlling colonisation density. Vaccine. 2019;37: 3953–3956. doi:10.1016/j.vaccine.2019.05.080

41. Dai Q, Dong Y, Wu J, Peng Q. Efficacy, immunogenicity, and safety of pneumococcal conjugate vaccine in children: a systematic review and meta-analysis. Front Pediatr. 2025;13. doi:10.3389/fped.2025.1652946

42. Higgins RA, Temple B, Dai VTT, Phan TV, Toan NT, Spry L, et al. Immunogenicity and impact on nasopharyngeal carriage of a single dose of PCV10 given to vietnamese children at 18 months of age. The Lancet Regional Health - Western Pacific. 2021;16: 100273. doi:10.1016/j.lanwpc.2021.100273

43. Hammitt LL, Ojal J, Bashraheil M, Morpeth SC, Karani A, Habib A, et al. Immunogenicity, Impact on Carriage and Reactogenicity of 10-Valent Pneumococcal Non-Typeable Haemophilus influenzae Protein D Conjugate Vaccine in Kenyan Children Aged 1–4 Years: A Randomized Controlled Trial. PLOS ONE. 2014;9: e85459. doi:10.1371/journal.pone.0085459

44. Whitney CG, Goldblatt D, O’Brien KL. Dosing Schedules for Pneumococcal Conjugate Vaccine. Pediatr Infect Dis J. 2014;33: S172–S181. doi:10.1097/INF.0000000000000076

45. Jayasinghe S, Chiu C, Quinn H, Menzies R, Gilmour R, McIntyre P. Effectiveness of 7- and 13-Valent Pneumococcal Conjugate Vaccines in a Schedule Without a Booster Dose: A 10-Year Observational Study. Clin Infect Dis. 2018;67: 367–374. doi:10.1093/cid/ciy129

46. Dunne EM, Pilishvili T, Adegbola RA. Assessing reduced-dose pneumococcal vaccine schedules in South Africa. The Lancet Infectious Diseases. 2020;20: 1355–1357. doi:10.1016/S1473-3099(20)30577-6

47. Kawade A, Dayma G, Apte A, Telang N, Satpute M, Pearce E, et al. Effect of reduced two-dose (1+1) schedule of 10 and 13-valent pneumococcal conjugate vaccines (SynflorixTM and Prevenar13TM)) on nasopharyngeal carriage and serotype-specific immune response in the first two years of life: Results from an open-labelled randomized controlled trial in Indian children. Vaccine. 2023;41: 3066–3079. doi:10.1016/j.vaccine.2023.04.008

48. Mackenzie GA, Osei I, Salaudeen R, Hossain I, Young B, Secka O, et al. A cluster-randomised, non-inferiority trial of the impact of a two-dose compared to three-dose schedule of pneumococcal conjugate vaccination in rural Gambia: the PVS trial. Trials. 2022;23: 71. doi:10.1186/s13063-021-05964-5

49. Quilty BJ, Toizumi M, Nguyen HAT, Le LT, Le HH, Iwasaki C, et al. Sustained and indirect effects of PCV10 reduced-dose schedules on pneumococcal carriage in Viet Nam: a long-term follow-up of a cluster-randomised controlled trial. The Lancet Infectious Diseases. 2026 [cited 17 June 2026]. doi:10.1016/S1473-3099(26)00172-6

50. Ladhani SN, Andrews N, Ramsay ME. Summary of evidence to reduce the two-dose infant priming schedule to a single dose of the 13-valent pneumococcal conjugate vaccine in the national immunisation programme in the UK. The Lancet Infectious Diseases. 2021;21: e93–e102. doi:10.1016/S1473-3099(20)30492-8

51. Report: Pneumococcal conjugate vaccine reduced dosing schedule: a systematic review and meta-analysis. Available: https://www.who.int/publications/m/item/report_who_sage_pcv_2025

52. O’Brien KL. When less is more: how many doses of PCV are enough? The Lancet Infectious Diseases. 2018;18: 127–128. doi:10.1016/S1473-3099(17)30684-9

53. Chen C-H, Janapatla RP, Su L-H, Li H-C, Kuo K-C, Chien C-C, et al. Incidence rates, emerging serotypes and genotypes, and antimicrobial susceptibility of pneumococcal disease in Taiwan: A multi-center clinical microbiological study after PCV13 implementation. Journal of Infection. 2022;84: 788–794. doi:10.1016/j.jinf.2022.04.022

54. Grant LR, Apodaca K, Deshpande L, Kimbrough JH, Hayford K, Yan Q, et al. Characterization of Streptococcus pneumoniae isolates obtained from the middle ear fluid of US children, 2011–2021. Front Pediatr. 2024;12: 1383748. doi:10.3389/fped.2024.1383748

55. Yokota S, Tsukamoto N, Sato T, Ohkoshi Y, Yamamoto S, Ogasawara N. Serotype replacement and an increase in non-encapsulated isolates among community-acquired infections of *Streptococcus pneumoniae* during post-vaccine era in Japan. IJID Regions. 2023;8: 105–110. doi:10.1016/j.ijregi.2023.07.002

56. Hanage WP, Auranen K, Syrjänen R, Herva E, Mäkelä PH, Kilpi T, et al. Ability of Pneumococcal Serotypes and Clones To Cause Acute Otitis Media: Implications for the Prevention of Otitis Media by Conjugate Vaccines. Infect Immun. 2004;72: 76–81. doi:10.1128/IAI.72.1.76-81.2004

57. Chen H-H, Hsu M-H, Wu T-L, Li H-C, Chen C-L, Janapatla RP, et al. Non-typeable *Streptococcus pneumoniae* infection in a medical center in Taiwan after wide use of pneumococcal conjugate vaccine. Journal of Microbiology, Immunology and Infection. 2020;53: 94–98. doi:10.1016/j.jmii.2018.04.001

58. Cohen R, Levy C, Ouldali N, Goldrey M, Béchet S, Bonacorsi S, et al. Invasive Disease Potential of Pneumococcal Serotypes in Children After PCV13 Implementation. Clin Infect Dis. 2021;72: 1453–1456. doi:10.1093/cid/ciaa917

59. de Miguel S, Pérez-Abeledo M, Ramos B, García L, Arce A, Martínez-Arce R, et al. Evolution of Antimicrobial Susceptibility to Penicillin in Invasive Strains of Streptococcus pneumoniae during 2007–2021 in Madrid, Spain. Antibiotics. 2023;12: 289. doi:10.3390/antibiotics12020289

60. Kawaguchiya M, Aung MS, Urushibara N, Ohashi N, Kobayashi N, Kudo K, et al. Molecular characterization and antimicrobial resistance of Streptococcus pneumoniae isolated from adult patients with invasive pneumococcal disease in northern Japan, 2017-2023. IJID Regions. 2025;16. doi:10.1016/j.ijregi.2025.100693

61. Méroc E, Fletcher MA, Hanquet G, Slack MPE, Baay M, Hayford K, et al. Systematic Literature Review of the Epidemiological Characteristics of Pneumococcal Disease Caused by the Additional Serotypes Covered by the 20-Valent Pneumococcal Conjugate Vaccine. Microorganisms. 2023;11: 1816. doi:10.3390/microorganisms11071816

62. Ntim OK, Kungu F, Narwortey DK, Donkor ES. Non-Typeable Streptococcus pneumoniae and Pneumococcal Diseases: A Systematic Review and Meta-Analysis. Tropical Medicine & International Health. 2026;n/a. doi:10.1111/tmi.70067

63. Temple B, Licciardi P. Is it time to reconsider how higher valency pneumococcal conjugate vaccines are evaluated? The Lancet Infectious Diseases. 2024;24: 228–229. doi:10.1016/S1473-3099(23)00634-5

64. CodeBlue. Pfizer’s 20-Valent Pneumococcal Conjugate Vaccine Now Approved In Malaysia. CodeBlue. 29 July 2025. Available: https://codeblue.galencentre.org/2025/07/pfizers-20-valent-pneumococcal-conjugate-vaccine-now-approved-in-malaysia/. Accessed 17 June 2026.

65. Bohari I. PELAKSANAAN PEMBERIAN 20-VALEN VAKSIN KONJUGAT PNEUMOKOKAL (PCV20) DALAM PROGRAM IMUNISASI PNEUMOKOKAL KANAK-KANAK. 2026.

66. Dunne EM, Struwig VA, Lowe W, Wilson CH, Perdrizet JE, Tamimi N, et al. Indirect Comparison of PCV20 Immunogenicity with PCV10 in Pediatric 3 + 1 and 2 + 1 Schedules. Infect Dis Ther. 2025;14: 1103–1117. doi:10.1007/s40121-025-01151-0

67. Janssens E, Flamaing J, Vandermeulen C, Peetermans WE, Desmet S, De Munter P. The 20-valent pneumococcal conjugate vaccine (PCV20): expected added value. Acta Clinica Belgica. 2022;78: 78–86. doi:10.1080/17843286.2022.2039865

68. Ngamprasertchai T, Ruenroengbun N, Kajeekul R. Immunogenicity and Safety of the Higher-Valent Pneumococcal Conjugate Vaccine vs the 13-Valent Pneumococcal Conjugate Vaccine in Older Adults: A Systematic Review and Meta-analysis of Randomized Controlled Trials. Open Forum Infect Dis. 2025;12: ofaf069. doi:10.1093/ofid/ofaf069

69. Al-Lahham A, Khanfar N, Albataina N, Al Shwayat R, Altwal R, Abulfeilat T, et al. Urban and Rural Disparities in Pneumococcal Carriage and Resistance in Jordanian Children, 2015–2019. Vaccines (Basel). 2021;9: 789. doi:10.3390/vaccines9070789

70. Bizuayehu HY, Kebede Y, Deressa M, Solomon E, Marami D, Karani A, et al. Pneumococcal carriage prevalence, serotype distribution, and vaccine coverage in Ethiopia 12 years after pneumococcal vaccine introduction. Vaccine. 2025;64: 127762. doi:10.1016/j.vaccine.2025.127762

71. Dunne EM, Choummanivong M, Neal EFG, Stanhope K, Nguyen CD, Xeuatvongsa A, et al. Factors associated with pneumococcal carriage and density in infants and young children in Laos PDR. PLOS ONE. 2019;14: e0224392. doi:10.1371/journal.pone.0224392

72. Lee C-C, Middaugh NA, Howie SRC, Ezzati M. Association of Secondhand Smoke Exposure with Pediatric Invasive Bacterial Disease and Bacterial Carriage: A Systematic Review and Meta-analysis. PLoS Med. 2010;7: e1000374. doi:10.1371/journal.pmed.1000374

73. Jacoby P, Carville KS, Hall G, Riley TV, Bowman J, Leach AJ, et al. Crowding and Other Strong Predictors of Upper Respiratory Tract Carriage of Otitis Media-related Bacteria in Australian Aboriginal and Non-Aboriginal Children. The Pediatric Infectious Disease Journal. 2011;30: 480. doi:10.1097/INF.0b013e318217dc6e

74. Parker AM, Jackson N, Awasthi S, Kim H, Alwan T, Wyllie AL, et al. Upper respiratory Streptococcus pneumoniae colonization among working-age adults with prevalent exposure to overcrowding. Microbiology Spectrum. 2024;12: e00879–24. doi:10.1128/spectrum.00879-24

75. Park B, Nizet V, Liu GY. Role of Staphylococcus aureus Catalase in Niche Competition against Streptococcus pneumoniae. Journal of Bacteriology. 2008;190: 2275–2278. doi:10.1128/jb.00006-08

76. Shak JR, Vidal JE, Klugman KP. Influence of bacterial interactions on pneumococcal colonization of the nasopharynx. Trends Microbiol. 2013;21: 129–135. doi:10.1016/j.tim.2012.11.005

77. Kovács E, Sahin-Tóth J, Tóthpál A, Linden M van der, Tirczka T, Dobay O. Co-carriage of Staphylococcus aureus, Streptococcus pneumoniae, Haemophilus influenzae and Moraxella catarrhalis among three different age categories of children in Hungary. PLOS ONE. 2020;15: e0229021. doi:10.1371/journal.pone.0229021

78. Tan ACH, Ramzi NH, Johari NA, Lai PK, Wong S, Chang XQ, et al. Co-carriage of respiratory tract bacterial pathogens among under-5 children with pneumococcal carriage in Peninsular Malaysia. BMC Infect Dis. 2026;26: 578. doi:10.1186/s12879-026-12800-1

79. Brotons P, Cisneros M, Pérez-Argüello A, Henares D, Lluansí A, Fernández de Sevilla M, et al. Pneumococcal nasopharyngeal carriage in children and adults self-confined at home during a COVID-19 national lockdown. PLoS One. 2024;19: e0315081. doi:10.1371/journal.pone.0315081

80. Kuo C-Y, Hwang K-P, Hsieh Y-C, Cheng C-H, Huang F-L, Shen Y-H, et al. Nasopharyngeal carriage of *Streptococcus pneumoniae* in Taiwan before and after the introduction of a conjugate vaccine. Vaccine. 2011;29: 5171–5177. doi:10.1016/j.vaccine.2011.05.034

81. Mills RO, Twum-Danso K, Owusu-Agyei S, Donkor ES. Epidemiology of pneumococcal carriage in children under five years of age in Accra, Ghana. Infectious Diseases. 2015;47: 326–331. doi:10.3109/00365548.2014.994185

82. Wang J, Qiu L, Bai S, Zhao W, Zhang A, Li J, et al. Prevalence and serotype distribution of nasopharyngeal carriage of Streptococcus pneumoniae among healthy children under 5 years of age in Hainan Province, China. Infect Dis Poverty. 2024;13: 7. doi:10.1186/s40249-024-01175-7

83. Dagan R, O’Brien KL. Modeling the Association between Pneumococcal Carriage and Child-Care Center Attendance. Clin Infect Dis. 2005;40: 1223–1226. doi:10.1086/428585

84. Manna S, McAuley J, Jacobson J, Nguyen CD, Ullah MdA, Sebina I, et al. Synergism and Antagonism of Bacterial-Viral Coinfection in the Upper Respiratory Tract. mSphere. 2022;7: e00984–21. doi:10.1128/msphere.00984-21

